# Association of Pluslife MiniDock MTB time-to-positivity with tuberculosis bacterial load: a diagnostic study in Indonesia

**DOI:** 10.64898/2026.09.01.26361863

**Authors:** Marcellus Korompis, Lara D. Veeken, Sri Hartati, Zuhaira H Fatma, Lidya Chaidir, Nina Eristiana, Todia Setiabudiawan, Jakko van Ingen, Reinout van Crevel, Philip C Hill, Rein M G J Houben, Bachti Alisjahbana, Raspati C Koesoemadinata

## Abstract

**Objectives:** The near point-of-care (nPOC) Pluslife MiniDock MTB (MiniDock) assay does not report semiquantitative values. We evaluated whether categorized MiniDock time-to-positivity (TTP) serves as a quantitative proxy for *Mycobacterium tuberculosis* (*Mtb*) bacterial load.

**Methods:** Presumptive tuberculosis (TB) patients enrolled across 27 health facilities in Indonesia were tested with sputum GeneXpert MTB/RIF Ultra (Xpert), MiniDock sputum swabs, and tongue swabs. Positive results were categorized using a median split at 13 minutes (≤13, 13-25, and 25 minute). MiniDock TTP categories were evaluated against Xpert semiquantitative grades and BACTEC MGIT 960 liquid culture TTP (days).

**Results:** Of 2974 presumptive TB participants tested with sputum Xpert, 426 (14.3%) were sputum Xpert-positive. MiniDock detected *Mtb* in 248/299 (83.0%) sputum and 263/382 (68.8%) tongue swab. Among 248 Minidock sputum-positive results, 99 (39.9%) turned positive ≤13 minutes, 107 (43.1%) between 13 and 25 minutes, and 42 (16.9%) at 25 minutes. Minidock TTP categories correlated with sputum and tongue swab semiquantitative results as well as with culture time to positivity (p<0.001).

**Conclusions:** MiniDock TTP categories (≤13, 13-25, 25 minutes) provide meaningful stratification which correlates with both Xpert semiquantitative and culture TTP. Time to Positivity from nPOC could thus serve as a proxy for bacterial burden and infectiousness, strongly increasing its utility for clinical care, public health and research.

## Introduction

In 2024, more than 8.3 million people were newly diagnosed with TB, but only around half of them were diagnosed with a WHO-recommended diagnostic test as their initial test^1^. Pluslife MiniDock MTB (MiniDock) is a novel near point-of-care (nPOC) assay that targets IS*6110* and *gyrB* gene by utilising loop-mediated isothermal amplification with RNase HII-mediated signal detection^2^. Unlike qPCR platforms, MiniDock does not generate cycle threshold (Ct) values. Assays such as GeneXpert MTB/RIF Ultra (Xpert) provide semiquantitative categories derived from Ct values, which have been shown to correlate with mycobacterial burden by smear and liquid culture time-to-positivity^3,4^, and are frequently used as measure of infectiousness^5,6^ or severity^7,8^. MiniDock results currently only report binary results (positive or negative) which have shown to correlate with sputum Xpert semiquantitative results, both for sputum and tongue swab samples^2^. One potential indicator of bacterial load in MiniDock is the time-to-positivity (TTP) output, defined as the time required for the RNase HII-mediated signal to reach the positivity threshold for both gene target. The test runs for 25 minutes, starts reporting after 10 minutes, turns positive within 25 minutes if both targets are detected, but only at 25 minutes if one target is detected (communication with manufacturer). In this study, we evaluated whether categorized classification of MiniDock TTP correlates with Xpert semiquantitative and culture TTP to serve as a proxy for bacterial load.

## Methods

### Study Population and sample collection

Data were derived from participants enrolled in the EVIDENT Indonesia (Evaluation and Demonstration of New Tuberculosis Diagnostics for Indonesia). Patients who were both positive for *Mycobacterium tuberculosis (Mtb)* by sputum Xpert and MiniDock from either sputum swab or tongue swab specimens were enrolled from 27 facilities in Bandung, Indonesia. We excluded patients if they were receiving TB preventive therapy, had TB disease within the last 2 years, had a severe illness, or were unable to produce spontaneous sputum. Sputum samples were used for Xpert, MiniDock testing, and liquid culture at facilities where culture testing was available. Not all diagnostic procedure were performed for every participant. Study protocol was approved by the Health Research Ethics Committee of Universitas Padjadjaran (Ref.No.616/UN6.KEP/EC/2024).

### Diagnostic Testing

Xpert sputum testing was performed according to manufacturer’s instructions^9^. Xpert results were categorized according to standard semiquantitative bacterial classifications of High, Medium, Low, Very Low and Trace^9^. MiniDock sputum swab and tongue swab testing were done according to manufacturer’s instructions. MiniDock results including TTP was recorded. TTP were converted to decimal minutes for analysis. In absence of manufacturer’s guidance or existing data, we divided TTP period based on median TTP in our study. Culture was available at 17 of the 27 participating facilities, using the BACTEC MGIT 960 (BD).

### Statistical Analysis

Data were analyzed in R 4.5.2 using *ggplot2* and *ggpubr*. Categorical associations between MiniDock TTP and Xpert semiquantitative grades were evaluated using Pearson’s Chi-squared test. Continuous liquid culture TTP days across MiniDock categories were compared using Kruskal-Wallis tests with post hoc Bonferroni-adjusted Wilcoxon rank-sum tests (*p*<0.05)

## Results

### MiniDock TTP categories and GeneXpert categories

We included 2974 participants with presumptive TB from EVIDENT that undergone sputum Xpert (Figure S1). Among those with a positive sputum MiniDock results before 25 minutes, 48.1% became positive at or before 13 minutes. Among positive sputum MiniDock results (n=248), 99 (39.9%) turned positive ≤13 minutes, 107 (43.1%) between >13 to <25 minutes, and 42 (16.9%) at 25 minutes. These categories correlated with sputum Xpert semiquantitative results (Figure 1A, *p*<0.001). Among tongue swab MiniDock results (n=263), 40 (15.2%) turned positive ≤13 minutes, 142 (54.0%) between >13 to <25 minutes, and 81 (30.8%) at 25 minutes. Similar as for sputum, these MiniDock TTP categories correlated with Xpert semiquantitative results (*p*<0.001) (Figure 1B) (Table 1). Associations were also observed across specimen types: shorter tongue swab MiniDock TTP was associated with higher sputum Xpert semiquantitative categories (*p*<0.001; Figure 1C), with a similar association between sputum MiniDock TTP and tongue swab Xpert semiquantitative categories (*p*=0.007; Figure 1D).

**Table 1.** Comparison of MiniDock TTP values (minutes) across different Xpert semiquantitative categories for sputum and TS samples. Data are reported as median [Interquartile Range] (number of positive samples). The minimum and maximum run time for the MiniDock assay is 10 to 25 minutes. Abbreviations: TS, Tongue Swab; SS, Sputum Swab; TTP, Time-to-Positive; NA, Not Applicable (no positive results obtained in this category).

| Xpert semiquantitative categories | Sputum Xpert |  |  |  | TS Xpert |  |  |  |
| --- | --- | --- | --- | --- | --- | --- | --- | --- |
|  | MiniDock Sputum Swab (N = 248) |  | MiniDock Tongue Swab (N=263) |  | MiniDock Sputum Swab (N = 106) |  | MiniDock Tongue Swab (N=63) |  |
|  | TTP in minutes (median [IQR]) (N total) | TTP Category (N per ≤13/ 13-25 / 25 mins) | TTP in minutes (median [IQR]) (N total) | TTP Category (N per ≤13/ 13-25 / 25 mins) | TTP in minutes (median [IQR]) (N total) | TTP Category (N per ≤13/ 13-25 / 25 mins) | TTP in minutes (median [IQR]) (N total) | TTP Category (N per ≤13/ 13-25 / 25 mins) |
| Trace | 25.0 [15.3-25.0] (n=12) | 1 / 3 / 8 | 25.0 [25.0-25.0] (n=6) | 0 / 1 / 5 | 25.0 [22.6-25.0] (n=12) | 0 / 3 / 9 | 15.6 [14.1-25.0] (n=7) | 0 / 4 / 3 |
| Very Low | 21.8 [14.6-25.0] (n=32) | 3 / 13 / 16 | 25.0 [14.6-25.0] (n=29) | 0 / 13 / 16 | 16.6 [14.6-25.0] (n=40) | 0 / 26 / 14 | 12.6 [12.6-13.6] (n=21) | 11 / 9 / 1 |
| Low | 14.6 [12.6-15.6] (n=71) | 19 / 38 / 14 | 15.6 [13.8-25.0] (n=66) | 10 / 31 / 25 | 14.6 [13.6-14.6] (n=50) | 11 / 36 / 3 | 12.6 [12.1-13.6] (n=31) | 19 / 11 / 1 |
| Medium | 12.6 [12.6-14.6] (n=59) | 30 / 28 / 1 | 14.6 [13.6-19.1] (n=72) | 13 / 43 / 16 | 12.6 [12.3-12.6] (n=4) | 4 / 0 / 0 | 12.6 [12.3-12.8] (n=4) | 3 / 1 / 0 |
| High | 12.6 [11.6-13.6] (n=74) | 46 / 25 / 3 | 14.6 [13.6-16.6] (n=90) | 17 / 54 / 19 | NA | NA | NA | NA |

**Figure 1.**
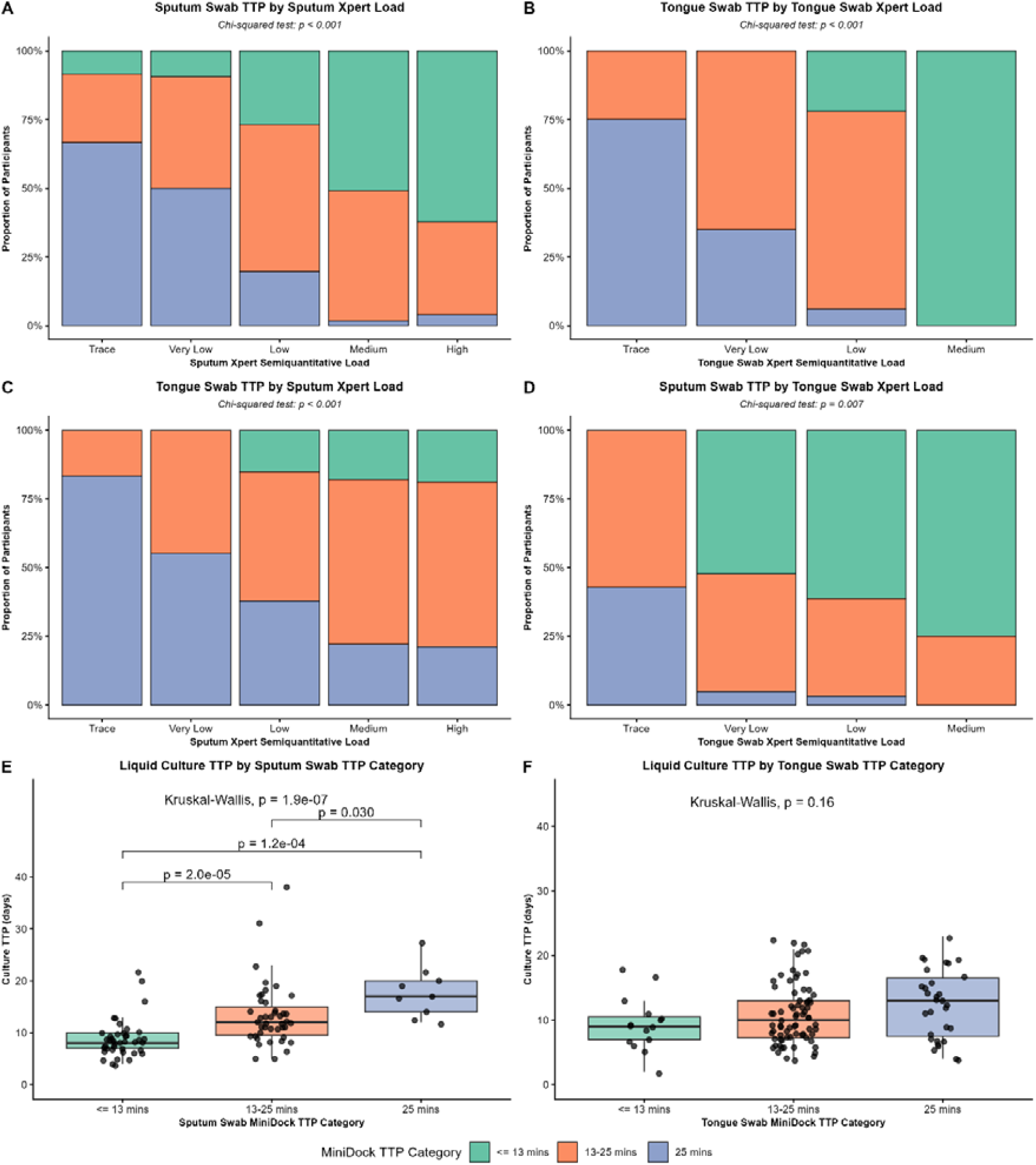
Association of Pluslife MiniDock TTP categories with GeneXpert Ultra semiquantitative and liquid culture TTP. (A–D) Bar charts displaying the proportion of positive participants (y-axis) in each MiniDock TTP category (≤13 mins (green), >13 to <25 mins (red), and 25 mins (blue)) across GeneXpert Ultra semiquantitative categories (x-axis). Same-compartment comparisons are shown for (A) SS TTP vs. sputum Xpert (n=248) and (B) TS TTP vs. TS Xpert (n=106). Cross-compartment comparisons are shown for (C) TS TTP vs. sputum Xpert (n=263) and (D) SS TTP vs. TS Xpert (n=63). p-values were derived from Pearson’s Chi-squared test. (E-F) Boxplots displaying liquid culture TTP in days (y-axis) across the MiniDock TTP categories (x-axis), with individual data points overlaid as jittered dots. Comparisons are shown for (E) liquid culture TTP by SS category (n=93) and (F) liquid culture TTP by TS category (n=120). Horizontal lines inside the boxes represent medians, and the bounds of the boxes represent the interquartile ranges. Brackets in panel E show pairwise Wilcoxon rank-sum test p-values adjusted for multiple testing using the Bonferroni method (non-significant comparisons are not shown). p-values are derived from Kruskal-Wallis and post-hoc Wilcoxon rank-sum test adjusted for multiple testing. Abbreviations: TTP, time-to-positivity; SQ, semiquantitative; SS, sputum swab; TS, tongue swab.

### MiniDock TTP categories and Liquid Culture TTP

Paired positive results of sputum culture were available for 93 individuals with positive sputum Minidock, and 120 with positive tongue swab MiniDock (Figure S1). Liquid culture TTP differed significantly across TTP categories for sputum MiniDock (Median [I QR], 8 [7-10], 12 [9.5-15], 17 [14-20] days), Figure 1E), but not for tongue swab MiniDock (Median [IQR], 9 [7-10.5], 10 [7.2-13], 13 [7.5-16.5] days, Figure 1F).

## Discussion

To our knowledge, this is the first study to formally establish MiniDock TTP as a quantitative proxy for TB bacterial load. While Mbuli et al.^10^ previously reported descriptive overall time-to-result figures across Xpert grades, our work advances substantially beyond this by introducing a 13-minute categorical threshold (≤13, 13-25, and 25 minutes). Importantly, we also demonstrated a clear stepwise relationship between sputum MiniDock TTP categories and liquid culture TTP.

MiniDock TTP categories associate with well-established Xpert semiquantitative values within both sputum and tongue swab specimens. Shorter MiniDock TTP (≤13 mins) correlated with High and Medium Xpert SQ results, while detection at the assay endpoint (25 mins) was associated with Trace and Very Low results. This demonstrates that MiniDock TTP provides quantitative information beyond the binary positive/negative result currently reported by the assay. Importantly, semiquantitative categories have been shown to predict infectiousness^11^ and disease severity^7^. Semiquantative results may also help stratify treatment duration, with a lower bacillary load allowing for shorter treatment ^7^.

Several limitations should be considered. First, not all participants underwent the same combination of diagnostic procedures. Second, Xpert is not specifically developed for tongue swab specimens, and its semiquantitative categories may not translate directly to tongue swab samples. Notably, both our data and Andama *et al*. showed that Xpert tongue swab may only reach the medium semiquantitative category^12^.

In conclusion, our results show that MiniDock TTP correlates with Xpert semiquantitative results and culture TTP and could thus serve as a proxy for bacterial burden and infectiousness, strongly increasing its utility for clinical care, public health and research.

## Supporting information

Figure S1. Participant enrolment flowchart. Positive Xpert results include all semiquantitative categories (High, Medium, Low, Very Low, and Trace).

## CRediT authorship contribution statement

Marcellus Korompis: Conceptualization, Methodology, Formal analysis, Data curation, Investigation, Writing (original draft), Writing (review & editing). Lara D. Veeken: Methodology, Formal analysis, Writing (review & editing). Sri Hartati: Data curation, Investigation, Writing (review & editing). Zuhaira H. Fatma: Data curation, Investigation, Methodology, Writing (review & editing). Lidya Chaidir: Conceptualization, Data curation, Formal analysis, Investigation, Methodology, Writing (review & editing). Nina Eristiana: Data curation, Investigation, Methodology, Writing (review & editing). Todia Setiabudiawan: Data curation, Investigation, Methodology, Writing (review & editing). Jakko van Ingen: Methodology, Writing (review & editing). Reinout van Crevel: Methodology, Writing (review & editing). Philip C. Hill: Conceptualization, Writing – review & editing. Rein M. G. J. Houben: Methodology, Writing (review & editing). Bachti Alisjahbana: Conceptualization, Resources, Supervision, Funding acquisition, Writing (review & editing). Raspati C. Koesoemadinata: Conceptualization, Funding acquisition, Resources, Supervision, Writing (review & editing).

## Transparency declaration

### Data availability statement

Data are available upon reasonable request.

## Acknowledgments

We thank all study participants and their families for their participation. We thank staff of Bandung District Health Office, Dr. H. A. Rotinsulu Lung Hospital, Dr. H. A. Rotinsulu Cibadak Primary Clinic, Puskesmas Ahmad Yani, Puskesmas Arcamanik, Puskesmas Astana Anyar, Puskesmas Babatan, Puskesmas Balai Kota, Puskesmas Caringin, Puskesmas Cetarip, Puskesmas Cigadung, Puskesmas Cijagra Baru, Puskesmas Cijagra Lama, Puskesmas Cipaku, Puskesmas Ciumbuleuit, Puskesmas Garuda, Puskesmas Griya Antapani, Puskesmas Gumuruh, Puskesmas Jajaway, Puskesmas Lio Genteng, Puskesmas Neglasari, Puskesmas Padasuka, Puskesmas Pagarsih, Puskesmas Pelindung Hewan, Puskesmas Puter, Puskesmas Sukahaji, Puskesmas Suryalaya, and Puskesmas Tamblong, for their support and participation in the study.

