## Supplementary material for "Association of Pluslife MiniDock MTB time-to-positivity with tuberculosis bacterial load: a diagnostic study in Indonesia": Figure S1. Participant enrolment flowchart. Positive Xpert results include all semiquantitative categories (High, Medium, Low, Very Low, and Trace).

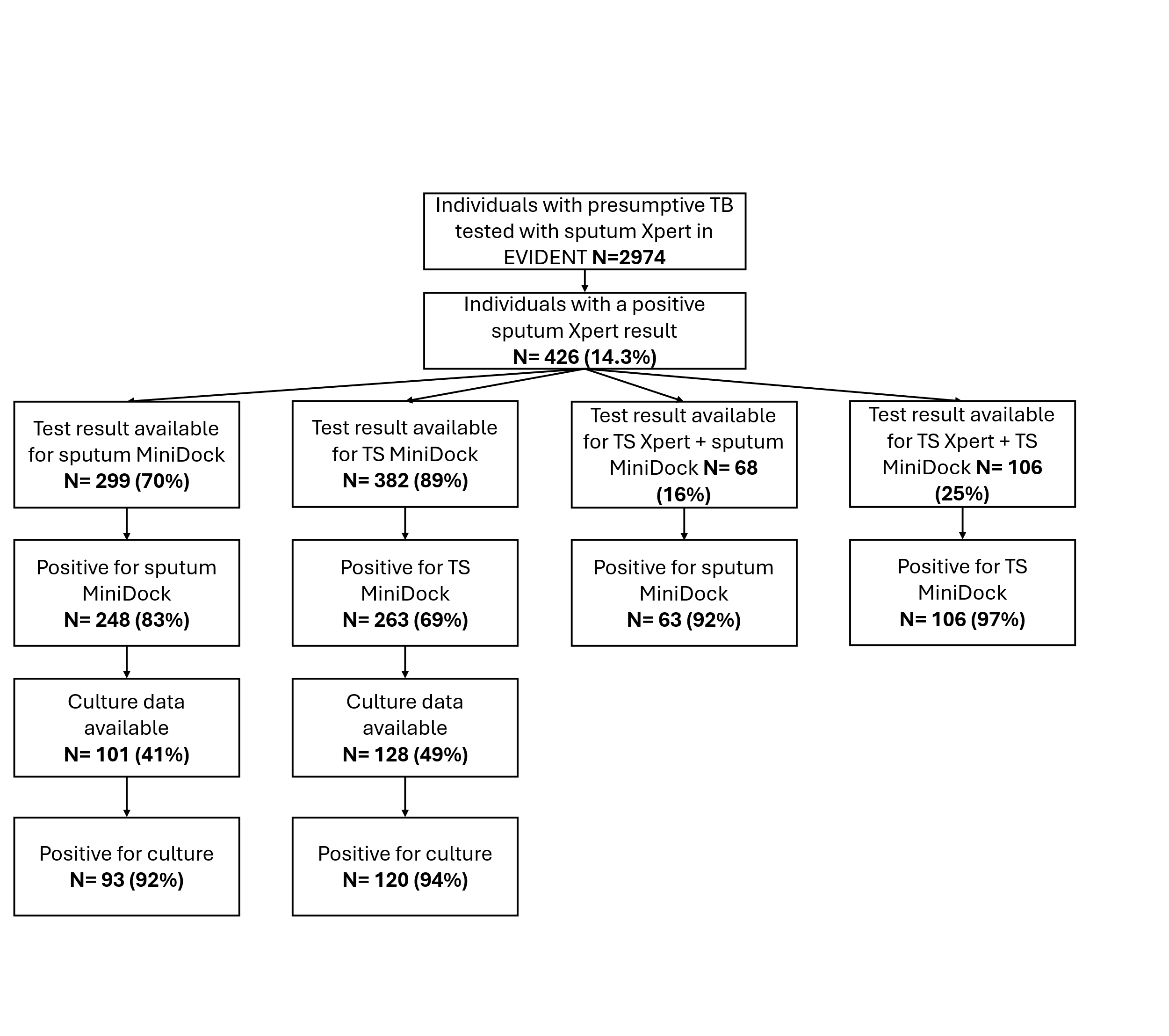

**Figure S1. Participant enrolment flowchart.** Positive Xpert results include all semiquantitative categories (High, Medium, Low, Very Low, and Trace). Participants without a positive MiniDock result had negative MiniDock test results. Culture data were unavailable for remaining participants because testing was not established in some participating community health facilities. Abbreviations: Xpert, GeneXpert MTB/RIF Ultra; TS, tongue swab
